# ASSESSMENT OF THE BACTERIOLOGICAL QUALITIES OF MEAT AND CONTACT SURFACES IN MARKETS IN ABIA STATE, NIGERIA

**DOI:** 10.1101/2023.10.04.23296550

**Authors:** Uchechukwu Olive Iwuagwu, Agwu Nkwa Amadi, Blessed Okwuchi Nworuh, Chimezie Christian Iwuala, David Chinaecherem Innocent, Michael Okwudiri Ikeanumba, Mary Onyinyechi Okorie

## Abstract

**Background:** Microbial contamination of meat comes from external sources during cutting, handling and processing of the meat. This study was carried out to assess the bacteriological qualities of meat and contact surfaces in markets in Abia State, Nigeria.

**Methods:** This research involved the use of a Hazard Analysis Critical Control Point (HACCP) checklist to investigate the sanitation and hygiene practices of meat sellers and a laboratory study of red and white meat, water and meat-contact surface samples. A total of 425 meat samples collected from 425 meat sellers from some randomly selected markets in Abia State were used for the study. There were also 20 water samples, 22 samples from table surfaces, 22 samples from knife surfaces and 14 samples from transport vehicles. The multistage simple random sampling technique through balloting was employed to determine communities/markets for the study. Samples for the study were collected and analyzed using standard microbiological techniques such as culturing and the bacteria were enumerated and identified using biochemical and chemical tests.

**Results:** The prevalent bacterial isolates include *Staphylococcus* sp (78.80%), *Bacillus* sp (73.17%), *Enterococcus* sp (64.00%), *Escherichia coli* (62.11%), *Salmonella* sp (62.11%), *Klebsiella* sp (51.29%), *Micrococcus sp* (44.94%) and *Campylobacter* sp (43.52%). SPSS analysis using the one way ANOVA showed no significant difference (P>0.05) in bacteria isolated from markets in the three Senatorial Zones of the State. *Staphylococcus* sp was isolated in 61.11% of the tables, 50.00% of vehicles, 41.67% of knives and 46.32% of water; *Salmonella* sp was isolated in 47.22% of the tables, 36.11% of vehicles, 30.56% of knives and 43.85% of water; *Bacillus* sp was isolated in 41.67% of the tables, 44.44% of vehicles, 33.33% of knives and 23.70% of water; *Campylobacter* sp was isolated in 27.78% of the tables, 25.00% of vehicles, 30.56% of knives and none in water. There was no significant difference (P>0.05) in bacteria isolated from the contact surfaces and water from the markets in the three zones of the State.

**Conclusion:** The bacteriological quality of meat in markets in Abia State could be said to be poor due to the isolation of Indicator bacteria such as *E, coli, Salmonella and Campylobacter* from the studied meat samples. The presence of *E. coli* in the studied meat samples is an indicator of feacal contamination and a red alert for the Public health sector. It is recommended that meat sellers undergo proper training and regularly update their knowledge of meat safety.

## 1.0 Introduction

Meat is a key component of food and it is rich in protein, mineral, vitamins and oil [1, 2]. Meat could be defined as the various tissues of animal origin and include beef from cattle, pork from pigs, mutton from sheep, poultry from chickens, ducks and turkey [1]. Fish, seafood, insects and snails are excluded here [3]. Meat is animal flesh that is eaten as food [4]. It is a good source of protein and amino acid (consisting of about 15 to 20 per cent of protein); iron; fat, zinc, B-vitamins, phosphorus etc. Meat supply protein which is of paramount importance as it is connected with the immune mechanism of the body; needed for building, repair and maintenance of body tissues; maintenance of osmotic pressure; and the synthesis of certain substances like antibodies, plasma proteins, haemoglobin, enzymes, hormones and coagulation factors [5].

However, this high nutrient, mineral and water contents of meat amongst other factors, predispose meat and meat products to microbial proliferations; resulting in their quick spoilage and contamination by microorganism [6]. With a high water content of about 75%, Fresh meats are among the most perishable foods [7]. Meat is one of the most perishable foods and is a good medium for microbial growth due its high nutrient and water contents, moderate pH and inherent chemical and enzymatic activities [8, 9, 10].

Thus, in order to ensure the wholesomeness, safety and quality of meat being sold to the public, various management procedures and guidelines for food and meat safety regulation nationally and internationally had been introduced. This is of paramount public health concern considering the continued global emergence and re-emergence of food borne diseases. Some of these internationally recommended meat/food safety protocols are the Codex Alimentarius Commission CAC - Good Hygiene Practices (GHPs) and Hazard Analysis Critical Control Point (HACCP) - based Standard Operating Procedures (SOPs). The HACCP approach is use to investigate the processes and procedures/management practices that contribute to bacterial contamination, growth and survival; and to identity points where control measures could be applied to prevent or eliminate the bacteriological hazards or reduce them to acceptable levels [11]. Meat handlers in Nigeria like their counterparts in other developing countries are yet to come to terms with these meat safety management protocols. Animals slaughtering and carcass handling in Nigeria also fall short of acceptable international standards; and that fresh meat sold to the public is contaminated from contact surfaces such as retail and slaughter slabs; dirty wheel barrows and car boots during transportation; openly displayed in the market and are examined with dirty hands by meat sellers and buyers with flies perching on them [12, 13].

Meat gets contaminated by microorganisms from external source during cutting, handling and processing of the meat - mainly from the skin and the intestinal tract of the animal [14]. Meat contamination could also occur during refrigeration if the proper cooling temperature is not maintained [15]. Some of the important bacteria that have been implicated in meat contamination and spoilage include: *Salmonella, Staphylococcus, Campylobacter, Escherichia coli, Enterobacter, Micrococcus, Bacillus, Clostridium, Streptomyces*, etc [16].

Food borne illness posed a significant public health challenges, and the prevention of food borne disease is an essential function of both public and environmental health [17]. Given the widespread impact of foodborne illness on people’s health, economies, and food systems amongst others; researchers from all over the world are committed to figuring out ways to increase food safety on a variety of levels. This has resulted in researches on food safety practices and food handling lately [18]. I intend to use HACCP-Good Hygiene Practices (GHPs) protocol for the assessment of meat hygiene and safety and then carry out bacteriological qualities assessment of meat and meat contact surfaces in markets in Abia State. It is generally believed that microbial contamination of meat comes from external sources during cutting, handling and processing of the meat. According to several publications, poor meat handling and management practices (in the storage, transportation and processing etc) at variance with internationally recognized standards as observed in some States in Nigeria have been implicated in meat contamination resulting in food poisoning and food borne diseases outbreaks; and other debilitating conditions such as kidney disorder (resulting from toxins produced by microorganisms in meat) [17, 18, 19, 20, 21, 22]. Some of these practices include non-adherence to internationally recommended standards such as the CAC - Good Hygiene Practices (GHPs) and HACCP - based Standard Operating Procedures (SOPs). It is reported that about 75 million cases of food poisoning and 5000 deaths occur annually in USA; with contaminated animal flesh accounting for 70% of the food poisoning [23, 24, 25]. Nigeria like other developing countries does not have accurate information on the prevalence and impact of food borne diseases; however, it is an established fact that diarrhea – the most common manifestation of food borne diseases is a major cause of sickness and death in the country. Contaminated meat/food is important cause of illness, disability and death globally; and food borne diseases impede socioeconomic development by straining health care systems; contributing to decrease in workers’ productivity; loss in school days; reduce family income as huge sums of money are spent on medical bills; causing pains and suffering and early death [22, 26]. These grave public health implications occasioned by increased mortality, morbidity and disability resulting from meat/food-borne diseases could be averted if internationally recognized food safety system such as HACCP-SOPs and GHP are incorporated in the meat management by meat handlers in Nigeria. I undertook this project due to the magnitude of the problem of poor hygiene practices and carcass handling in Nigeria which fall short of acceptable international standards with the resultant contamination of fresh meat sold to the public and the attendant problems. The general objective of this study is to assess the bacteriological qualities of meat and contact surfaces in markets in Abia State, Nigeria.

## 2.0 Methods

### 2.1 Study Design

This research design was an experimental study involving laboratory tests/analysis to assess the bacteriological qualities of meat and meat contact surfaces in markets in Abia State.

### 2.2 Study Setting

Abia state was created from part of Imo state in 27^th^ August 1991. The geographical coordinates of Abia state is 5.4309°N 7.5247°E. As at the 2006 census, the population of Abia state was put at 2,833,999. Its capital city is Umuahia and the major commercial city is Aba. English is widely spoken and serves as the official language in governance and business. Christianity is the predominant religion of Abia people. Abia state has 3 senatorial zones with 17 Local Government Areas (LGAs). The senatorial zones are Abia Central, Abia North and Abia South. The LGAs include: Aba North, Aba South, Arochukwu, Bende, Ikwuano, Isiala Ngwa North, Isiala Ngwa South, Isuikwuato, Obi Ngwa, Ohafia, Osisioma Ngwa, Ugwunagbo, Ukwa East, Ukwa West, Umuahia North, Umuahia South and Umu Nneochi. Figure 1.0 shows the 3 senatorial zones and the LGAs in each zone of Abia state.

**Figure 1:**
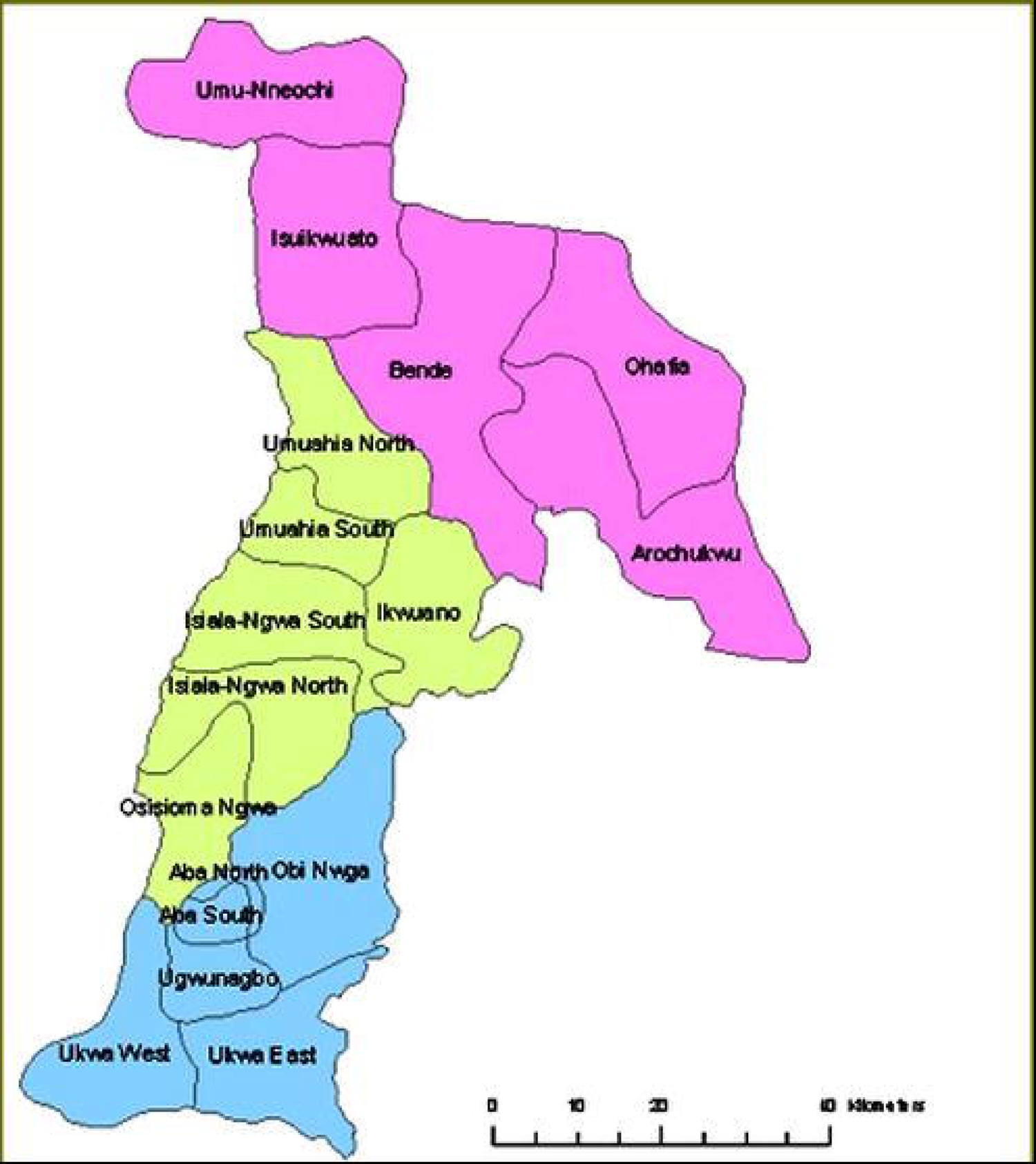
Geographyical Map of the Study Area-Abia State showing the three (3) Senatorial Zones and LGAs (Source: Nigerian Muse, 2010)

### 2.3 Study Population

Meat handlers include meat handlers in abattoirs/ slaughter houses; meat handlers in the markers (meat sellers) and meat handlers in transit from abattoirs to markets.

The study population here is meat (red and white) sellers in markets in Abia State, Nigeria. According to the information from the meat sellers Associations in Abia State, there are about three thousand one hundred (3100) meat sellers across the various markets in Abia State. Ten (10) Local Government Areas (LGAs) out of the Seventeen (17) LGAs from the three Senatorial Zones in Abia State were randomly through balloting selected for this study.

### 2.4 Sample Size and Sampling Technique

#### 2.4.1 Sample Size

The sample size calculation of the population of meat sellers in the markets for this study was determined using Taro Yamane (1967) formula:

N = N/1+N(e)^2^

Where n=sample size; N=Population size; e=Level of precision (5%) n= 3100/1+3100 x (0.05)^2^

= 3100/1+3100 x .0025

=4250/1 + 7.75

=3100/8.75

= 354.28 approximately 354

Adding 20% to account for attrition, then the 20% of 354 = 0.20 x 354 = 70.85 approximately

71

Therefore, the total sample size for this study is 354 + 71 = **425 meat sellers**

#### 2.4.2 Sampling Technique

A Multi stage simple random sampling technique was adopted for this study.

##### 2.4.2.1 Selection of LGAs, Markets

A simple random sampling using balloting was used for the selection of ten (10) out of the seventeen (17) Local Government Areas (LGAs) in Abia State for the study thereby giving every LGA in Abia State an equal chance of selection by the researcher. Thereafter, through balloting, markets were selected from enumerated major markets in the selected LGAs and communities for sampling.

##### 2.4.2.2 Selection of Respondents

The respondents together with the meat samples were randomly selected through balloting whereby all respondents present at the time of study who picked even numbers were selected until the minimum sample size of the study was obtained.

Thus, a total of 425 samples of meat and meat sellers were randomly selected from markets in ten (10) LGAs in Abia State, Nigeria was used for the study.

The sampled markets in Aba, Umuahia and Ohafia Senatorial Zones have a total number of 340, 250 and 100 meat sellers respectively out of which 200, 160 and 65 randomly selected meat sellers were drawn/participated in this study from the three senatorial zones respectively.

Table 1.0 below shows the distribution of participating meat sellers in the sampled markets according to the Senatorial Zones in Abia State

**Table 1.0:**
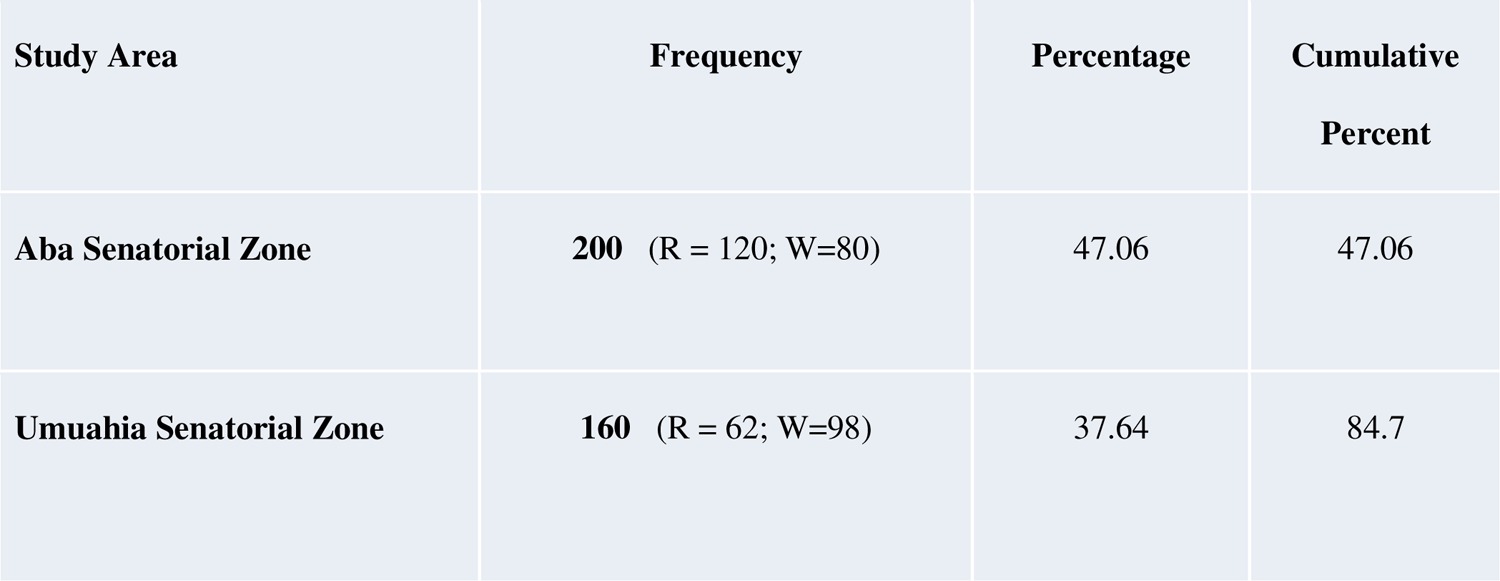
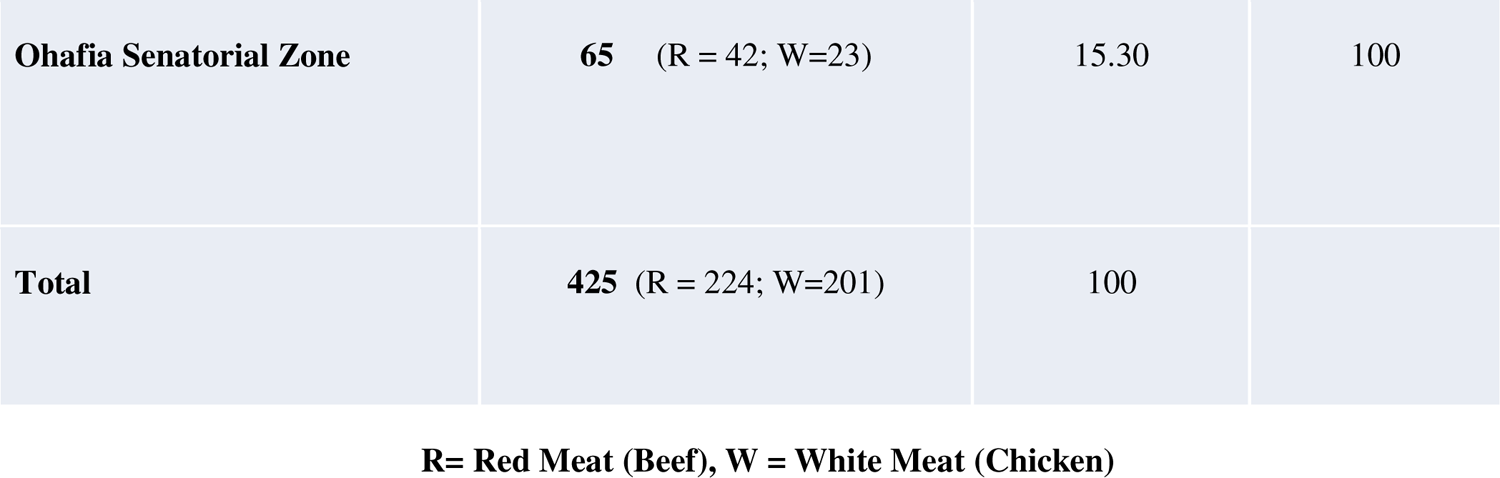
Distribution of participating meat sellers/ meat samples in the sampled markets according to the Senatorial Zones in Abia State.

| **Study Area** | **Frequency** | **Percentage** | **Cumulative**<br>**Percent** |
| --- | --- | --- | --- |
| **Aba Senatorial Zone** | **200** (R = 120; W=80) | 47.06 | 47.06 |
| **Umuahia Senatorial Zone** | **160** (R = 62; W=98) | 37.64 | 84.7 |
| **Ohafia Senatorial Zone** | **65** (R = 42; W=23) | 15.30 | 100 |
| **Total** | **425** (R = 224; W=201) | 100 |  |

**R= Red Meat (Beef), W = White Meat (Chicken)**

### 2.5 Inclusion and Exclusion Criteria

All meat sellers in the markets (both male and females from the ages of 18 years and above) who practice their trade in Abia State; and gave their consent for the study were part of this research work. Meat sellers/handlers who did not give an informed consent to be part of the study were excluded.

### 2.6 Instrument for Data Collection

The instrument for data collection were questionnaire and Laboratory equipment for the assessment of the bacteriological qualities of meat samples.

#### 2.6.1 Laboratory equipment and materials for the bacteriological assessment

##### Glass Wares

1. Test tube
2. Pipettes (1ml, 2mls, 5mls, 10mls)
3. Conical flasks (100mls. 250mls, 500mls, 1000mls)
4. Beaker (100mls, 250mls, 500mls, 1000mls)
5. Glass spreader
6. Glass slides
7. Cover slips
8. Bijou bottles
9. Measuring cylinder (250mls, 500mls, 1000mls)
10. Volumetric flask (100mls, 250mls, 500mls, 1000mls)
11. Centrifuge tubes
12. Durham tubes **Personal Protective Equipment**
13. Hand gloves
14. Face mask
15. Nose mask
16. Laboratory coat **Reagents and Chemicals**
17. Alcohol
18. Acetone
19. Kovacs reagents
20. Alpha naphthol
21. Indole reagent **Media and Diluents**
22. Distilled water
23. Peptone water
24. Normal Saline Solution
25. Grams reagents
26. Methyl red
27. Bromothymol blue
28. Methylene blue
29. MR-VP Broth
30. Glucose
31. Sucrose
32. Lactose
33. Mannitol
34. Maltose
35. Fructose
36. Salmonella Shigella Agar
37. Eosin Methylene Blue Agar
38. MacConkey Agar
39. Nutrient Agar
40. Nutrient Broth
41. Campylobacter Blood Free Agar
42. Salmonella Shigella Agar
43. Sodium Chloride
44. Mannitol Salt Agar
45. Simmon’s Citrate agar **Equipment for Laboratory analysis**
46. Colony counter
47. Microscope
48. pH meter
49. Water Bath
50. Autoclave
51. Hot Air Oven
52. Centrifuge
53. Refrigerator
54. Laboratory blender
55. Weighing balance (electronic and manual)
56. Digital Meat Thermometer **Other materials**
57. Cotton wool
58. Masking tape
59. Markers
60. Blotting paper
61. Wire loop
62. Mounting needle/inoculating needle
63. Gas cylinder
64. Cold box

### 2.7 Procedure for Samples (Data) Collection/preparation and analysis

#### 2.7.1 Collection of Samples

Four hundred and twenty-five meat samples comprising 224 red meat-beef (120 from Aba zone, 62 from Umuahia zone and 42 from Ohafia zone) and 201 white meat-chicken (80 from Aba zone, 98 from Umuahia zone and 23 from the Ohafia zone) were collected from markets in Abia State. Collections of the meat samples were in sterile containers and collected samples were transported in an ice packed cooler to the laboratory. Samples were also taken from the water sources (20 samples) in the market and contact surfaces of the meat handlers which included tables (22 samples), knives (22 samples) and transport vehicles(14 samples). Samples from contact surfaces were collected using sterile specimen sponges wetted with 10ml of buffered peptone water (Oxoid) from sterile Whirl-Pak bags (Sponge-Bag, PBI-International) using a template of 100cm^2^ surface area. Sponging within the selected area consisted of 5 passes vertically (up and down was considered as one pass) and then 5 passes horizontally (side to side was considered one pass). The sponge was placed into a Stomacher bag, labelled and delivered in a cold box to the laboratory within 4 hours. All collected samples were properly labelled and taken to the Environmental Health Laboratory, College of Health Sciences, Abia State University Aba for analysis. Copies of the questionnaires were administered to the meat sellers with the help of Field Assistants who have already been trained for that purpose.

#### 2.7.2 Preparation of Media and Diluents

All bacteriological media were prepared according to manufacturer’s specification. Nutrient agar was used in the isolation of heterotrophic bacteria, MacConkey Agar for faecal coliform bacteria, Eosin Methylene Blue Agar for *Escherichia coli*, Campylobacter Agar for *Campylobacter* species, Mannitol Salt Agar strictly for *Staphylococcus aureus* and Salmonella Shigella Agar for the isolation of *Salmonella* and *Shigella* species). Physiological saline used as diluents was prepared by dissolving 9.8 g of sodium chloride in 1000ml of distilled and dispensed in 90 ml and 9ml portions. Both diluents and media were sterilized in an autoclave at 121^0^C for 15 minutes.

#### 2.7.3 Sample analysis and Tests: Preparation of Samples and Inoculation

Ten (10) grams of meat sample was macerated in a sterile laboratory blender containing 90 ml of sterile peptone water. Ten-fold dilution method was used by transferring 1 ml from each tube until the required dilution was obtained. Aliquot portion (0.1ml) of appropriate dilution was inoculated into the pre-sterilized and surface dried medium. Inocula were spread evenly to ensure uniform and countable colonies and plated on different types of media for microbial growth and enumeration. Plates were incubated at 28°C for 48 hours for heterotrophic bacteria.

Test tubes containing swabs were shaken on a vortex mixer for 30 seconds for uniform distribution of bacteria. Tenfold serial dilution of all samples was prepared using sterile normal saline solution (NSS) 0.1ml of each sample was pipetted into agar plate and incubated at 37°C for 42-48 hours for total viable bacteria count.

For Total Aerobic Mesophilic Count (TAMC), on an agar media plate, 0.1ml of each sample was pipetted and spread. Inoculated plate was incubated at 32°C for 48-72 hours.

For Total coliforms and fecal Coliform Count, 0.1ml of each sample was pipetted and spread on Violet red Bile agar. Inoculated plate was incubated at 32°C for 18-24 hours to determine the total coliforms; and at 44.5°C for 18-24 hours to determine the fecal coliform.

For Enterobacteriaceae Count, 0.1ml of each sample was pipetted and spread on MacConkey agar supplemented with glucose. Inoculated plate was incubated at 35°C for 24 hours. All reddish purple/pink colonies were counted as members of the Enterobacteriaceae.

For Aerobic spore former bacterial count, meat sample suspension was first heated at 80°C in the water bath for ten minutes to kill the vegetative cells. Then, 0.1ml of each sample was pipetted and spread on plate count agar (PCA) plate. Inoculated plate was incubated at 35°C for 36-72 hours.

#### 2.7.4 Determination of Microbial Population

After incubation, plates with colonies between 30 and 300 were counted using Colony counter and the result were expressed as colony forming units per gram (CFU/g) to obtain total population.

#### 2.7.5 Characterization and Identification of Microbial Isolates

After incubation of the various inoculated plates, the predominant bacterial colonies were picked randomly from countable plates and inoculated into test tubes containing about 5ml nutrient broth. The bacterial culture were purified by repeated streak plating and characterization. The predominant bacterial isolates were characterized based on cultural (colonial), microscopic and biochemical methods with reference to standard manuals. The identities of the isolates were cross-matched with reference to standard manuals for the identification of bacteria [2].

### Microscopic Characterization

#### i. Gram Staining Test

The Gram staining technique was used for the bacterial isolates as described by Cheesbrough [2]. A smear of the isolate was made on grease free glass slide with a drop of water and allowed to dry. The smear was fixed by mild heating, flooded with crystal violet and allowed to stand for 30 seconds. The crystal violet was rinsed off with water. Lugol’s iodine was added and allowed to stand for 30 seconds. This was washed off with water and acid alcohol, till discoloration. It was counter stained with Safranin for 10 seconds and rinsed with water. The wet slide was allowed to air dry. A drop of oil immersion was added on the slide and viewed using X100 objective lens of the microscope.

#### ii. Spore Staining Test

The spore stain was used to confirm the presence of spores when indicated in the Gram stain. Isolates were heat fixed on a slide and flooded with 5% malachite green. It was steamed for 3 minutes (without allowing it to boil), dried and cooled. It was then rinsed off and stained with Safranin for 30 seconds. This was rinsed, dried with filter paper and viewed under the microscope using oil immersion tens. The positive spores showed green while the negative cells were stained pink.

#### iii. Motility Test

This test was used to determine the motility of bacteria isolated. The test was carried out on a semi-solid agar medium in which motile bacteria swarm and gave a diffuse spreading growth. The medium was dispensed into test tubes, sterilized and allow to set in an upright position. It was then inoculated using an inoculation needle by stabbing it into the medium in the test tube. This was incubated at 37°C for 24 hours. Diffuse growth from the straight line of inoculation was recorded as positive result [2].

### Biochemical Characterization of Bacteria Isolates

Microorganisms that were not identified by the colonial and microscopic characteristics were further subjected to few biochemical tests described by Cheesbrough [2].

#### i. Catalase Test

The enzyme catalase is present in most cytochrome containing aerobic and facultative anaerobic bacteria. Catalase has one of the highest turnover numbers of all enzymes such that one molecule of catalase can convert millions of molecules of hydrogen peroxide to water and oxygen in a second. Catalase activity can be detected by adding the substrate HDOD to an appropriately incubated (18-24 hours) tryptic soy agar slant culture. Organisms which produce the enzyme breakdown the hydrogen and the resulting OD production produces bubbles in the reagent drop indicating a positive test. Organisms lacking the cytochrome system also lack the catalase enzyme and are unable to breakdown peroxide into OD and water and are catalase negative.

#### ii. Coagulase Test

Coagulase is an enzyme that clots blood plasma by a mechanism that is similar to normal clotting. The coagulase test identifies whether an organism produces this exoenzyme. This enzyme clots the plasma component of blood. The only significant disease-causing bacteria of humans that produce coagulase are *Staphylococcus aureus.* Thus, this enzyme is a good indicator of *S. aureus.* In the test, the sample is added to rabbit plasma and held at 37DC for a specified period of time. Formation of clot within four hours is indicated as positive result and indicative of a virulent *Staphylococcus aureus* strain. The absence of coagulation after 24 hours of incubation is a negative result indicative of a virulent strain.

#### iii. Oxidase Test

Oxidase test is an important differential procedure that should be performed on all gram-negative bacteria for their rapid identification. The test depends on the ability of certain bacteria to produce indophenol blue from the oxidation of dimethyl-p-phenylenediamine and α-naphthol. This method uses N, N-dimethyl-p-phenylenediamine oxalate in which all *Staphlococci* are oxidase negative. In the presence of the enzyme cytochrome oxidase (gram negative bacteria) the N, N-dimethhyl-p-phenylenediamine oxalate and α-naphthol react to indophenol blue. *Pseudomonas aeruginosa* is an oxidase positive organism.

#### iv. Sugar Fermentation/Oxidation

This test is used to differentiate between bacteria groups that oxidize carbohydrate such as members of Enterobacteriaceae. One milliliter (1ml) of 10% glucose, maltose, lactose, fructose, mannitol, and sucrose were separately under aseptic conditions transferred into duplicate tubes containing 9ml of sterile Hugh and Leifson’s medium to obtain a final concentration of 1% of each of sugar. The tubes were stab-inoculated in duplicates while two un-inoculated tubes serve as control. Vaseline was used to cover one set of the duplicate tubes, one control to discourage oxidative utilization of sugar. All tubes were incubated at 37°C for 48 hours. After the incubation, they were observed for acid production in the culture. Yellow coloration indicates acid production in the open tubes only suggesting oxidative utilization of the sugar while acid production in the sealed tubes suggests a fermentative reaction.

#### v. Hydrogen Sulphide Production (H_2_S) Test

The test isolates were aseptically inoculated into a tube containing triple sugar iron agar started by stabbing the agar to the bottom and streaking the surface of the slant. The inoculated tube was incubated at 37°C for 72 hours and was examined daily. Black precipitation and yellow coloration was checked for. Black precipitate indicates H_2_S production and yellow coloration for sucrose, lactose and glucose fermentation.

#### vi. Urease Test

Urease Agar slant in McCartney bottle was inoculated with the bacteria isolate at 30°C for 4 hours and then overnight. A pink color in the medium indicated a positive result.

#### vii. IMViC Test

This test consists of four different test; they are Indole production, Methyl-Red test, Voges Proskaeur test and Citrate utilization test. This test is specifically designed to determine the physiological properties of microorganism. They are especially useful in the differentiation of Gram-negative intestinal bacilli, particularly *Escherichia coli* and the *Enterobacter-Klebsiella* group.

#### viii. Indole Test

This test demonstrates the ability of certain bacteria to decompose the amino acid-Tryptophan to Indole. The bacteria isolates were inoculated into the medium and incubated at 37°C for 48 hours. At the end of incubation period, 3 drops of kovac’s reagents was added and then shaken. A red color ring at the interface of the medium denotes a positive result. Methyl red and Voges-Proskaeur test must be considered together since they are physiologically related. Opposite test is usually obtained from the MR and VP test, that is, MR+, VP-, or MR-, VP+.

Methyl red test was performed to demonstrate the capacity of different organisms to produce acid from the fermentation of sugar (dextrose). Methyl-red positive organisms produce a red coloration when five drops of methyl-red indicator is added into 48 hour old MR-VP broth culture.

The Voges-Proskaeur test demonstrates the ability of organisms to produce acetone from glucose metabolism. Some organisms metabolize glucose to produce pyruvic acid which is further broken down to yield Butane-diol and acetyl-methyl carbinol as an intermediate product. Into one milliliter of the culture, one milliliter of six percent alcoholic solution of alpha-naphtol was added to one milliliter of 16% KOH and allowed to stand for 15-20 minutes. Development of a red to pink color was a positive test.

#### ix. Citrate Utilization Test

This is one of the several techniques used to assist in the identification of Enterobacteria. Principle of the test is based on the ability of an organism to use citrate as its only source of carbon. The test was carried out using Simmon’s citrate agar. The slopes of the media were prepared in bijou bottles as recommended by the manufacturers. A sterile straight wire was used to the slope with a saline suspension of the test organisms before stabbing the butt. The bottles were incubated at 37°C for 48 hours. Bright blue colors in the medium means positive test while no change in color of medium indicates negative citrate test [2]

### 2.8 Method of Data Analysis

The data from this research work was collated manually by the Researcher; and then entered into the computer by a statistician. The Statistical Package for the Social Sciences (SPSS) software (version 20) was used in the analysis of the data. Results were expressed in percentages, frequencies, tables. One-way ANOVA and the independent sample T-test was used to test the hypotheses at 95% confidence interval and 0.05 Level of significance.

### 2.9 Ethical clearance/ Informed Consent

An informed consent was gotten from all meat handlers who participated in the study. The purpose of the research was explained to each respondent and verbal informed consent obtained from them before inclusion into the study. Also, anonymity of the respondents was assured and ensured.

## 3.0 Results

A total of four hundred and twenty-five (425) meat samples comprising 224 red meat-beef (120 from Aba zone, 62 from Umuahia zone and 42 from Ohafia zone) and 201 white meat-chicken (80 from Aba zone, 98 from Umuahia zone and 23 from Ohafia zone) collected from four hundred and twenty-five (425) meat sellers from markets in Abia State were used for this study. There were also twenty (20) water samples, twenty-two (22) samples from table surfaces, twenty-two (22) samples from knife surfaces and fourteen (14) samples from transport vehicles. The results of Data collected and analyized are presented in the tables below

### 3.1 Bacteriological Qualities Of Meat Samples and Meat Contact Surfaces

A total of 425 meat samples comprising 224 red meat-beef (120 from Aba zone, 62 from Umuahia zone and 42 from Ohafia zone) and 201 white meat-chicken (80 from Aba zone, 98 from Umuahia zone and 23 from Ohafia zone) were collected and analysed. There were also twenty (20) water samples, twenty-two (22) samples from table surfaces, twenty-two (22) samples from knife surfaces and fourteen (14) samples from transport vehicles.

The results of the predominant bacterial isolates of the meat samples and the meat contact surface from markets in Abia State were as presented in the tables below:

### 3.2 Bacteria isolated from the meat samples (red and white meat) from markets in Abia State

The result of the Bacteria isolated from the 425 meat samples (red and white meat) from markets in Abia State is presented in the table 4.6 below.

Table 2.0 showed that out of the total of 425 meat samples collected and analyzed, *Staphylococcus sp*. was isolated in 335 (78.80%) of the meat samples; *Escherichia coli*, 264 (62.11%); *Micrococcus* sp., 191 (44.94%); *Salmonella* sp., 264 (62.11%); *Bacillus* sp., 311 (73.17%); *Campylobacter* sp., 185 (43.52%); *Klebsiella* sp., 218 (51.29%); *Enterococcus* sp., 272 (64.00%); *Shigella* sp., 106 (24.94%); *Pseudomonas* sp., 64 (15.05%); *Enterobacter* sp., 161 (37.88%)

**Table 2.0.**
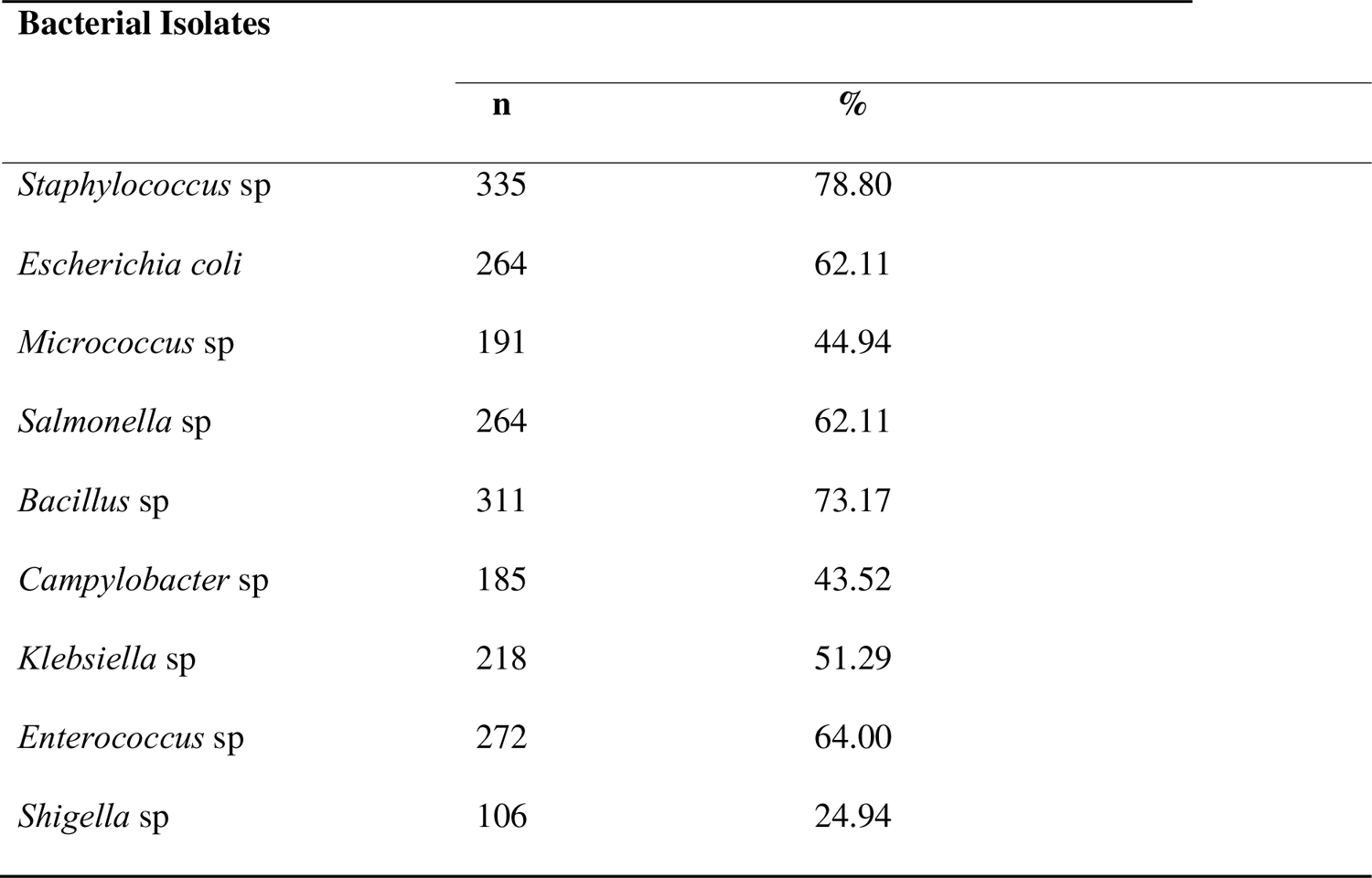
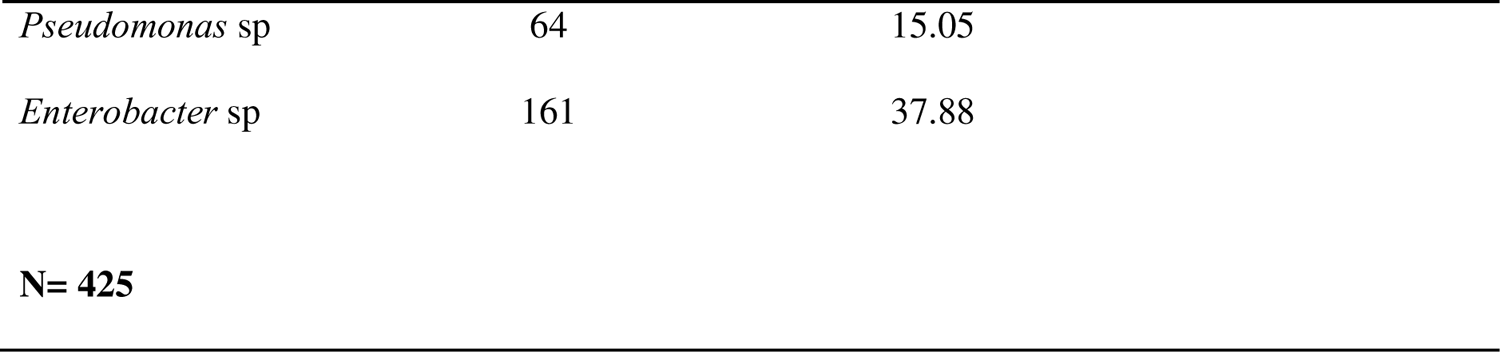
Bacteria isolated from the meat samples (red and white meat) from markets in Abia State.

| **Bacterial Isolates** |  |  |
| --- | --- | --- |
|  | **n** | **%** |
| *Staphylococcus sp* | 335 | 78.80 |
| *Escherichia coli* | 264 | 62.11 |
| *Micrococcus sp* | 191 | 44.94 |
| *Salmonella sp* | 264 | 62.11 |
| *Bacillus sp* | 311 | 73.17 |
| *Campylobacter sp* | 185 | 43.52 |
| *Klebsiella sp* | 218 | 51.29 |
| *Enterococcus sp* | 272 | 64.00 |
| *Shigella sp* | 106 | 24.94 |
| *Pseudomonas* sp | 64 | 15.05 |
| *Enterobacter* sp | 161 | 37.88 |
| **N= 425** |  |  |

### 3.3 Bacteria isolated from red meat samples from markets in Abia State

The result of the Bacteria isolated from red meat samples from markets in Abia State is presented in the table 3.0 below. A total of 224 red meat samples were collected and analysed (120 from Aba zone, 62 from Umuahia zone and 42 from Ohafia zone)

**Table 3.0.** Bacteria isolated from red meat samples in markets in Abia state.

| **Bacterial Isolates** |  |  |
| --- | --- | --- |
|  | **n** | **%** |
| *Staphylococcus* sp | 175 | 78.12 |
| *Escherichia coli* | 139 | 62.05 |
| *Micrococcus* sp | 100 | 44.64 |
| *Salmonella* sp | 138 | 61.60 |
| *Bacillus* sp | 167 | 74.55 |
| *Campylobacter* sp | 99 | 44.19 |
| *Klebsiella* sp | 116 | 51.78 |
| *Enterococcus* sp | 142 | 63.39 |
| *Shigella* sp | 55 | 24.55 |
| *Pseudomonas* sp | 34 | 15.17 |
| *Enterobacter* sp | 84 | 37.50 |
| **N= 224** |  |  |

Table 3.0 showed that out of the 224 red meat sampled, *Staphylococcus* sp was isolated in 175 (78.12%) of the red meat samples; *Escherichia coli*, 139 (62.05%); *Micrococcus* sp., 100 (44.64%); *Salmonella* sp., 138 (61.60%); *Bacillus* sp., 167 (74.55%); *Campylobacter* sp., 99 (44.19%); *Klebsiella* sp., 116 (51.78%); *Enterococcus* sp., 142 (63.39%); *Shigella* sp., 55 (24.55%); *Pseudomonas* sp., 34 (15.17%); *Enterobacter* sp., 84 (37.50%)

### 3.4 Comparison of bacteria isolated from red meat samples from markets in the Three Senatorial zones in Abia State

The result of the comparison of the Bacteria isolated from 224 red meat-beef samples(120 from Aba zone, 62 from Umuahia zone and 42 from Ohafia zone) from markets in the three Senatorial Zones in Abia State is presented in the table 4.0 below.

**Table 4.0:** Comparison of bacteria isolated from red meat samples from markets in the Senatorial Zones in Abia state.

| Bacterial Isolates | Umuahia |  | Aba |  | Ohafia |  |
| --- | --- | --- | --- | --- | --- | --- |
|  | n | % | n | % | n | % |
| *Staphylococcus* sp | 50 | 81.09 | 93 | 77.18 | 32 | 77.11 |
| *Escherichia coli* | 40 | 64.12 | 72 | 60.00 | 27 | 65.06 |
| *Micrococcus* sp | 30 | 48.10 | 52 | 43.55 | 18 | 43.55 |
| *Salmonella* sp | 40 | 65.28 | 71 | 59.19 | 27 | 65.14 |
| *Bacillus* sp | 46 | 73.43 | 91 | 76.23 | 30 | 71.28 |
| *Campylobacter* sp | 25 | 40.58 | 53 | 43.84 | 21 | 49.84 |
| *Klebsiella* sp | 32 | 51.40 | 60 | 50.10 | 24 | 58.25 |
| *Enterococcus* sp | 43 | 68.81 | 72 | 60.05 | 27 | 64.33 |
| *Shigella* sp | 16 | 25.66 | 30 | 24.68 | 9 | 22.61 |
| *Pseudomonas* sp | 9 | 14.92 | 17 | 13.91 | 8 | 18.16 |
| *Enterobacter* sp | 25 | 39.70 | 45 | 37.60 | 14 | 33.89 |

Table 4.0 showed that *Staphylococcus* sp was isolated in 81.09% of the red meat in Umuahia, 77.18% in Aba and 77.11% in Ohafia; *Escherichia coli*, 64.12% in Umuahia, 60.00% in Aba and 65.06% in Ohafia; *Micrococcus* sp, 48.10% in Umuahia, 43.55% in Aba and 43.55% in Ohafia; *Salmonella* sp, 65.28% in Umuahia, 59.19% in Aba and 65.14% in Ohafia; *Bacillus* sp, 73.43% in Umuahia, 76.23% in Aba and 71.28% in Ohafia; *Campylobacter* sp, 40.58% in Umuahia, 43.84% in Aba and 49.84% in Ohafia; *Klebsiella* sp., 51.40% in Umuahia, 50.10% in Aba and 58.25% in Ohafia; *Enterococcus* sp., 68.81% in Umuahia, 60.05% in Aba and 64.33% in Ohafia; *Shigella* sp., 25.66% in Umuahia, 24.68% in Aba and 22.61% in Ohafia; *Pseudomonas* sp., 14.92% in Umuahia, 13.91% in Aba and 18.16% in Ohafia; *Enterobacter* sp., 39.70% in Umuahia, 37.60% in Aba and 33.89% in Ohafia.

SPSS analysis using the one-way ANOVA showed that there was no statistically significant difference between bacteria isolated from red meat samples from markets in the Senatorial Zones in Abia State [p = 0.7678) > 0.05].

### 3.5 Bacteria isolated from white meat samples from markets in Abia State

The result of the Bacteria isolated from white meat-chicken samples from markets in Abia State is presented in the table 5.0 below. A total of 201 white meat (chicken) samples were collected and analysed (80 from Aba zone, 98 from Umuahia zone and 23 from Ohafia zone)

**Table 5.0:** Bacteria isolated from white meat samples in markets in Abia State.

| **Bacterial Isolates** | **n** | **%** |
| --- | --- | --- |
| *Staphylococcus* sp | 160 | 79.60 |
| *Escherichia coli* | 125 | 62.18 |
| *Micrococcus* sp | 91 | 45.27 |
| *Salmonella* sp | 126 | 62.68 |
| *Bacillus* sp | 144 | 71.64 |
| *Campylobacter* sp | 86 | 42.78 |
| *Klebsiella* sp | 102 | 50.74 |
| *Enterococcus* sp | 130 | 64.67 |
| *Shigella* sp | 51 | 25.37 |
| *Pseudomonas* sp | 30 | 14.92 |
| *Enterobacter* sp | 77 | 38.30 |
| **N= 201** |  |  |

Table 5.0 showed that out of the total 201 white meat sampled, *Staphylococcus* sp was isolated in 160 (79.60%) of the white meat samples; *Escherichia coli*, 125 (62.18%); *Micrococcus* sp., 91 (45.27%); *Salmonella* sp., 126 (62.68%); *Bacillus* sp., 144 (71.64%); *Campylobacter* sp., 86 (42.78%); *Klebsiella* sp., 102 (50.74%); *Enterococcus* sp., 130 (64.67%); *Shigella* sp., 51 (25.37%); *Pseudomonas* sp., 30 (14.92%); *Enterobacter* sp., 77 (38.30%);

### 3.6 Comparison of bacteria isolated from white meat samples from markets in the Three Senatorial zones in Abia State

The result of the comparison of the Bacteria isolated from 201white meat-chicken samples(80 from Aba zone, 92 from Umuahia zone and 23 from Ohafia zone) from markets in the three Senatorial Zones in Abia State is presented in the table 6.0 below.

**Table 6.0:** Comparison of bacteria isolated from white meat samples from markets in the Senatorial Zones in Abia state.

| Bacterial Isolates | Umuahia |  | Aba |  | Ohafia |  |
| --- | --- | --- | --- | --- | --- | --- |
|  | n | % | n | % | n | % |
| *Staphylococcus* spp | 80 | 81.18 | 62 | 77.17 | 18 | 76.12 |
| *Escherichia coli* | 63 | 64.12 | 48 | 60.05 | 14 | 62.53 |
| *Micrococcus* sp | 47 | 48.10 | 35 | 43.52 | 9 | 42.50 |
| *Salmonella* sp | 64 | 65.22 | 47 | 59.11 | 15 | 64.17 |
| *Bacillus* sp | 72 | 73.44 | 56 | 70.28 | 16 | 71.45 |
| *Campylobacter* sp | 40 | 40.53 | 35 | 43.88 | 11 | 47.60 |
| *Klebsiella* sp | 50 | 51.47 | 40 | 50.10 | 12 | 51.13 |
| *Enterococcus* sp | 67 | 68.86 | 48 | 60.04 | 15 | 64.34 |
| *Shigella* sp | 25 | 25.69 | 20 | 24.66 | 6 | 27.70 |
| *Pseudomonas* sp | 15 | 14.90 | 11 | 13.98 | 4 | 15.41 |
| *Enterobacter* sp | 39 | 39.73 | 30 | 37.67 | 8 | 36.51 |
| **ANOVA P value = 0.19** |  |  |  |  |  |  |
| **Decision = NS** |  |  |  |  |  |  |

NS\* - Not Significant; S\* - Significant

Table 6.0: showed that *Staphylococcus* sp was isolated in 81.18% of the white meat in Umuahia, 77.17% in Aba and 76.12% in Ohafia; *Escherichia coli*, 64.12% in Umuahia, 60.05% in Aba and 62.53% in Ohafia; *Micrococcus* sp, 48.10% in Umuahia, 43.52% in Aba and 42.50% in Ohafia; *Salmonella* sp, 65.22% in Umuahia, 59.11% in Aba and 64.17% in Ohafia; *Bacillus* sp, 73.44% in Umuahia, 70.28% in Aba and 71.45% in Ohafia; *Campylobacter* sp, 40.53% in Umuahia, 43.88% in Aba and 47.60% in Ohafia; *Klebsiella* sp., 51.47% in Umuahia, 50.10% in Aba and 51.13% in Ohafia; *Enterococcus* sp., 68.86% in Umuahia, 60.04% in Aba and 64.34% in Ohafia; *Shigella* sp., 25.69% in Umuahia, 24.66% in Aba and 27.70% in Ohafia; *Pseudomonas* sp., 14.90% in Umuahia, 13.98% in Aba and 15.41% in Ohafia; *Enterobacter* sp., 39.73% in Umuahia, 37.67% in Aba and 36.51% in Ohafia.

SPSS analysis using the one-way ANOVA showed no significant difference [P(0.19)>0.05] between bacteria isolated from white meat samples from markets in the Senatorial Zones in Abia state.

### 3.7 Comparison of bacteria isolated from red and white meat samples from markets in Abia State

The result of the comparison of the Bacteria isolated from the 224 red meat and 201white meat samples from markets in Abia State is presented in the table 7.0 below.

**Table 7.0:**
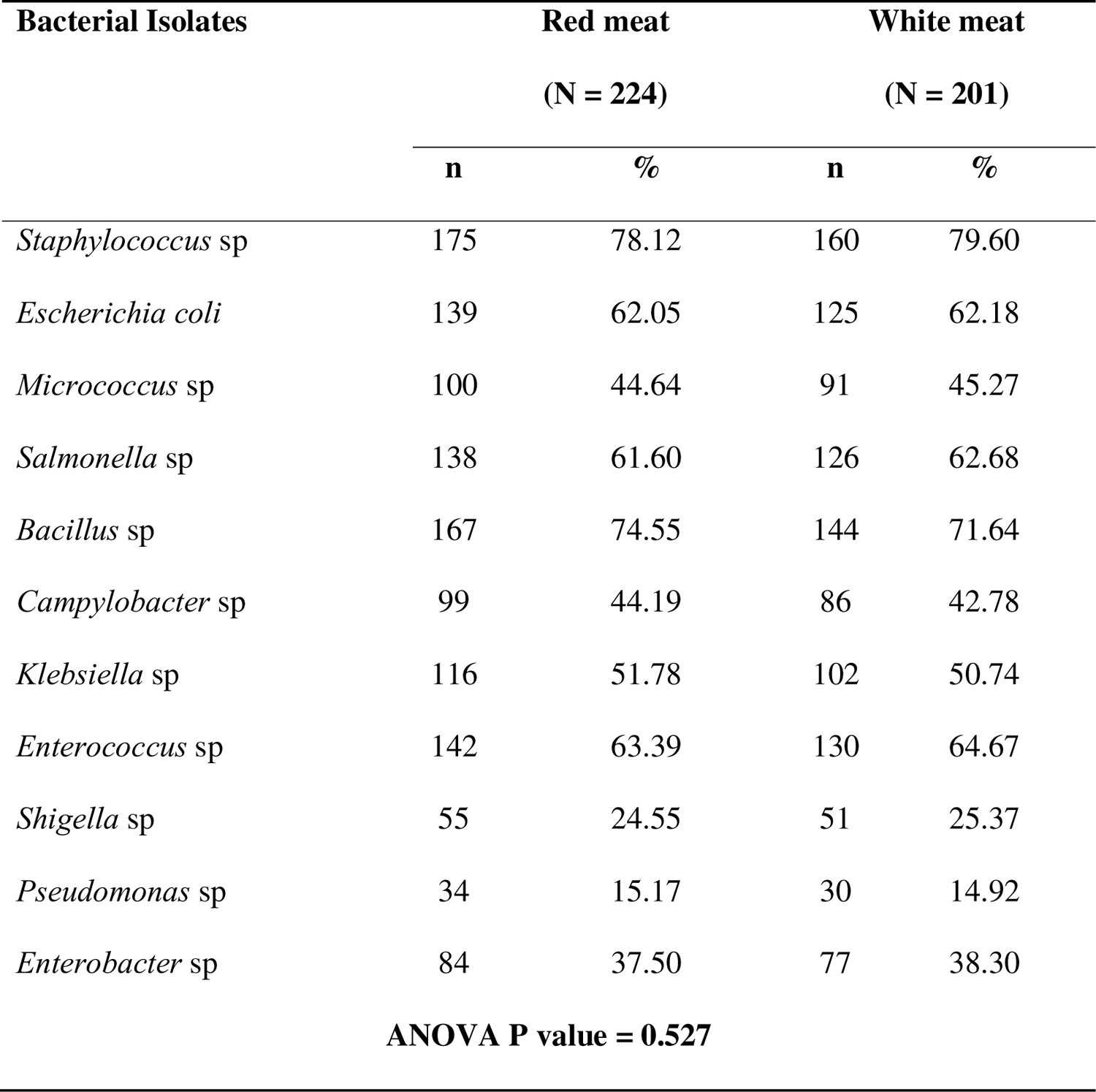
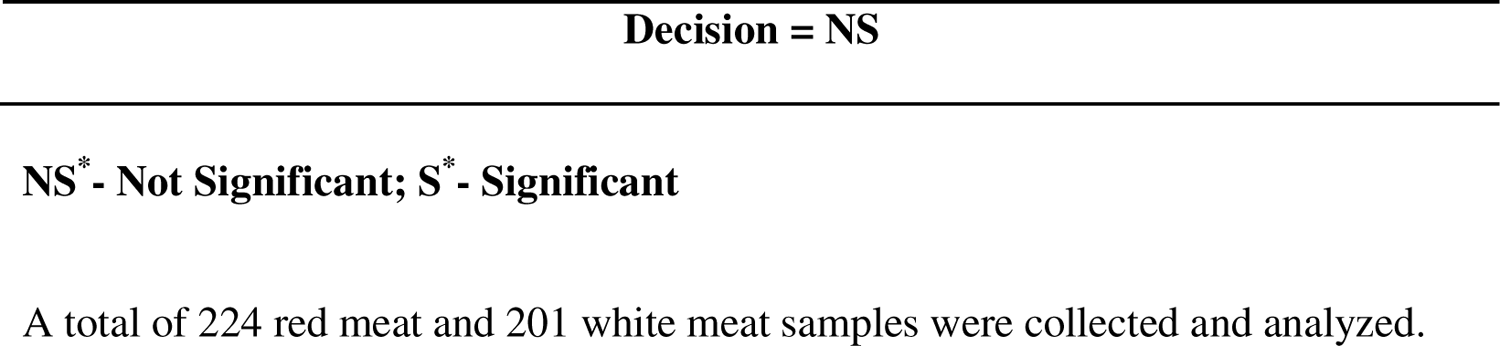
Comparison of Bacteria isolated from red and white meat samples from Markets in Abia state.

| Bacterial Isolates | Red meat<br>(N = 224) |  | White meat<br>(N = 201) |  |
| --- | --- | --- | --- | --- |
|  | n | % | n | % |
| *Staphylococcus* sp | 175 | 78.12 | 160 | 79.60 |
| *Escherichia coli* | 139 | 62.05 | 125 | 62.18 |
| *Micrococcus* sp | 100 | 44.64 | 91 | 45.27 |
| *Salmonella* sp | 138 | 61.60 | 126 | 62.68 |
| *Bacillus* sp | 167 | 74.55 | 144 | 71.64 |
| *Campylobacter* sp | 99 | 44.19 | 86 | 42.78 |
| *Klebsiella* sp | 116 | 51.78 | 102 | 50.74 |
| *Enterococcus* sp | 142 | 63.39 | 130 | 64.67 |
| *Shigella* sp | 55 | 24.55 | 51 | 25.37 |
| *Pseudomonas* sp | 34 | 15.17 | 30 | 14.92 |
| *Enterobacter* sp | 84 | 37.50 | 77 | 38.30 |
| **ANOVA P value = 0.527** |  |  |  |  |

NS\* - Not Significant; S\* - Significant
A total of 224 red meat and 201 white meat samples were collected and analyzed.

Table 7.0 showed that out of the total 224 red and 201 white meat sampled, *Staphylococcus* sp was isolated in 175 (78.12%) of the red meat samples and 160 (79.60%) of the white meat samples; *Escherichia coli*, 139 (62.05%) red meat and 125 (62.18%) white; *Micrococcus* sp., 100 (44.64%) red meat and 91 (45.27%) white; *Salmonella* sp., 138 (61.60%) red meat and 126 (62.68%) white; *Bacillus* sp., 167 (74.55%) red meat and 144 (71.64%); *Campylobacter* sp., 99 (44.19%) red meat and 86 (42.78%) white; *Klebsiella* sp., 116 (51.78%) red meat and 102 (50.74%) white; *Enterococcus* sp., 142 (63.39%) red meat and 130 (64.67%) white; *Shigella* sp., 55 (24.55%) red meat and 51 (25.37%) white; *Pseudomonas* sp., 34 (15.17%) red meat and 30 (14.92%) white; *Enterobacter* sp., 84 (37.50%) red meat and 77 (38.30%) white. SPSS analysis using the one-way ANOVA showed no significant difference [P(0.527)>0.05] in bacteria isolated from red and white meat samples from markets in Abia State.

### 3.8 Comparison of bacteria isolated found on meat contact surface samples - tables, knives, water, and transport Vehicles in use in markets in Abia State

The result of the comparison of the Bacteria isolated from 78 meat contact surface samples (comprised of 22 table surfaces, 22 knives surfaces, 14 transport vehicles and 20 water samples) from markets in Abia State is presented in the table 8.0 below.

**Table 8.0:** Comparison of bacterial isolates found on tables, knives, water, and transport.

| Bacteria | Table |  | Vehicle |  | Knife |  | Water |  |
| --- | --- | --- | --- | --- | --- | --- | --- | --- |
|  | n | % | n | % | n | % | n | % |
| *Staphylococcus* sp | 13 | 61.11 | 7 | 50.00 | 9 | 41.67 | 9 | 46.32 |
| *Escherichia coli* | 0 | 0.00 | 0 | 0.00 | 0 | 0.00 | 0 | 0.00 |
| *Micrococcus* sp | 0 | 0.00 | 0 | 0.00 | 0 | 0.00 | 0 | 0.00 |
| *Salmonella* sp | 10 | 47.22 | 5 | 36.11 | 7 | 30.56 | 9 | 43.85 |
| *Bacillus* sp | 9 | 41.67 | 6 | 44.44 | 7 | 33.33 | 5 | 23.70 |
| *Campylobacter* sp | 6 | 27.78 | 4 | 25.00 | 7 | 30.56 | 0 | 0.00 |
| *Klebsiella* sp | 0 | 0.00 | 0 | 0.00 | 0 | 0.00 | 0 | 0.00 |
| *Enterococcus* sp | 0 | 0.00 | 0 | 0.00 | 0 | 0.00 | 0 | 0.00 |
| *Shigella* sp | 0 | 0.00 | 0 | 0.00 | 0 | 0.00 | 0 | 0.00 |
| *Pseudomonas* sp | 0 | 0.00 | 0 | 0.00 | 0 | 0.00 | 0 | 0.00 |
| *Enterobacter* sp | 0 | 0.00 | 0 | 0.00 | 0 | 0.00 | 0 | 0.00 |
| **P value = 0.8100** |  |  |  |  |  |  |  |  |
| **Decision = NS** |  |  |  |  |  |  |  |  |

**NS\* - Not Significant; S\* - Significant**

Table 8.0 below showed that *Staphylococcus* sp was isolated in 13 (61.11%) of the 22 samples from tables, 7 (50.00%) of the 14 samples from vehicles, 9 (41.67%) of the 22 samples from knives, and 9 (46.32%) of the 20 water samples; *Salmonella* sp, in 10 (47.22%) of the tables, 5 (36.11%) of vehicles, 7 (30.56%) of knives and 9 (43.85%) of water samples; *Bacillus* sp., in 9 (41.67%) of the tables, 6 (44.44%) of vehicles, 7 (33.33%) of knives and 5 (23.70%) of water samples; *Campylobacter* sp., in 6 (27.78%) of the tables, 4 (25.00%) of vehicles, 7 (30.56%) of knives and none in water. No other bacteria were isolated from the samples.

SPSS analysis using the one-way ANOVA showed that there was no statistically significant difference between the bacterial isolates found on tables, knives, water, and transport vehicles in use in markets in Abia State (p = 0.8100).

## 4.0 Discussion

The results of this study revealed the presence of various bacterial isolates in the meat samples (both red and white meat), with *Staphylococcus* sp, *Bacillus* sp, *Escherichia coli*, *Enterococcus* sp, *Salmonella* sp, *Klebsiella* sp, *Micrococcus sp*, and *Campylobacter* sp being the prevalent isolates. These findings are of great concern as they indicate the potential for bacterial contamination in the meat sold in the markets. *Staphylococcus* sp was the most prevalent isolate, present in a high percentage (78.80%) of the meat samples. This bacterium is known to be commonly associated with human skin and can be transferred to meat during handling and processing, highlighting the significance of proper hygiene practices among meat handlers. Similarly, *Escherichia coli*, a common indicator of fecal contamination, was found in a substantial proportion (62.11%) of the meat samples, suggesting possible contamination from improper slaughter and processing practices. Other bacterial isolates, such as *Micrococcus* sp, *Salmonella* sp, *Bacillus* sp, and *Campylobacte*r sp, were also detected at varying rates. The presence of these organisms on the surface of meat samples and the contact surfaces, such as tables, vehicles, and knives, indicates potential fecal and environmental contamination. The poor personal hygiene and sanitation practices among meat sellers/handlers as observed in this study could have contributed to the contamination of the meat. Most of the predominant bacteria in the meat contact surfaces such as *Staphylococcus* sp., *Salmonella* sp., *Bacillus* sp., and *Campylobacter* sp. were also predominant in the meat samples suggesting a possible cross-contamination of the meat carcasses from the contact surfaces. This contamination can occur during various stages, from slaughter to transportation and display of meat in the markets. The findings of this study are in agreement with previous studies by other researchers including Gutema *et al.,* [27] who reported and linked the isolation of *Salmonella* sp from contaminated chicken meat to poor sanitary and sanitation conditions; Shimelis, *et al.* [28] who in their studies isolated *E. coli* and *Salmonella* species as the common bacterial isolates from beef at selected slaughter houses and attributed their sources of contamination to include equipment, transport vehicle, cutting board and worker’s hand.

Also, the statistical analysis showed no significant difference in the bacteria isolated from the various markets (P>0.05), suggesting that the prevalence of these bacterial isolates is consistent across the studied markets. This finding raises concerns about the overall hygiene and sanitation practices in the meat markets, as the presence of these bacteria on meat surfaces can pose significant health risks to consumers. The high prevalence of these bacterial isolates underscores the importance of implementing stringent hygiene and sanitation measures in meat handling and processing as already suggested by previous researchers including Azuamah *et al.,* [19] and Tesson *et al.,* [29].

Meat sellers/handlers should be trained on proper hygiene practices, including handwashing, wearing gloves and aprons, and ensuring the cleanliness of equipment and contact surfaces. Additionally, market authorities should enforce regulations and conduct regular inspections to ensure compliance with hygiene and sanitation standards. The findings of this study are consistent with previous research on meat contamination and highlight the need for continuous monitoring and improvement of meat handling practices to ensure the safety and quality of meat products. By addressing the issues of bacterial contamination in meat markets, public health risks can be minimized, and consumers can have greater confidence in the safety of the meat they purchase and consume.

## 5.0 CONCLUSION

In conclusion, the evaluation of the bacteriological quality of meat and contact surfaces in markets in Abia State, Nigeria, revealed an alarmingly low level of hygiene and sanitation practises among meat vendors and handlers. The presence of Salmonella spp. and Escherichia coli indicator bacteria on meat samples and contact surfaces, such as tables, vehicles, and knives, demonstrates the potential for faecal and environmental contamination. This contamination is likely the result of poor personal hygiene and suboptimal meat processing procedures during slaughter, dressing, and other stages of production. The findings highlight the urgent need for comprehensive education and awareness campaigns to improve meat handling practises among individuals involved in the meat industry in Abia State, thereby protecting public health and ensuring the safety of meat products offered for sale.

### Contribution to Knowledge

This study has added to existing knowledge that meat sellers/handlers in Abia states, Nigeria failed to meet the basic standards of personal hygiene and sanitation practices during the handling of meat in the sampled markets; and this could have lead to the compromised the bacterial qualities of the meat being sold. This study successfully isolated, identified and documented the predominant bacterial isolates on meat sold in markets in Abia State as *Staphylococcus sp., Escherichia sp., Salmonella sp., Bacillus sp., Entercoccus sp*. and *Campylobacter sp.*; while *Staphylococcus sp., Salmonella sp., Bacillus sp., and Campylobacter sp.* were predominant bacteria on the meat contact surfaces. This study has also showed that the contamination of meat could have come from external sources through cross contamination of the meat carcasses from the various contact surfaces during handling. The bacteriological quality of the meat sold across markets in Abia State indicated that there is a Process hygiene criteria failure in meat handling which does not called for the withdrawal of meat being sold to the public but for corrective measures to be put in place to correct reoccurrence. Meat sellers/handlers in Abia State should be enlightened/trained on proper meat handling standards of operation.

### Recommendations

Meat handlers are advised to undergo proper training and regular update on their knowledge of meat safety especially on proper sanitation and good personal hygiene practices. Meat handlers should be educated on the need to comply with standard operation procedures for the handling of meat such as the wearing of aprons and gloves, proper temperature control, and improved means of transportation. Government and Non-govermental agencies should fabricate a prototype of a customized cold chain chest for storage and transportation of meat; as well as for display of meat in the markets.

### Ethics Approval and consent to Participate

Not Applicable

### Consent to Publish

Not applicable

### Availability of Data and Materials

The Data set from the study are available to the corresponding author upon request.

## Competing Interests

Authors have declared that they have no competing interests

## Funding

No funds were received for this study

## Data Availability

All data produced in the present study are available upon reasonable request to the authors

## Acknowledgements

Not applicable

## Notes

### Competing Interest Statement

The authors have declared no competing interest.

### Funding Statement

This study did not receive any funding.

